# Automated and partially-automated contact tracing: a rapid systematic review to inform the control of COVID-19

**DOI:** 10.1101/2020.05.27.20114447

**Authors:** Isobel Braithwaite, Tom Callender, Miriam Bullock, Robert W Aldridge

**Author notes:** Correspondence:* Dr Isobel Braithwaite, Institute of Health Informatics, University College London, 222 Euston Road, London, NW1 2DA.

## Abstract

**Background:** Automated or partially-automated contact tracing tools are being deployed by many countries to contain SARS-CoV-2; however, the evidence base for their use is not well-established.

**Methods:** We undertook a rapid systematic review of automated or partially-automated contact tracing, registered with PROSPERO (CRD42020179822). We searched PubMed, EMBASE, OVID Global Health, EBSCO COVID Portal, Cochrane Library, medRxiv, bioRxiv, arXiv and Google Advanced for articles relevant to COVID-19, SARS, MERS, influenza or Ebola from 1/1/2000-14/4/2020. Two authors reviewed all full-text manuscripts. One reviewer extracted data using a pre-piloted form; a second independently verified extracted data. Primary outcomes were the number or proportion of contacts (and/or subsequent cases) identified; secondary outcomes were indicators of outbreak control, app/tool uptake, resource use, cost-effectiveness and lessons learnt. The Effective Public Health Practice Project tool or CHEERS checklist were used in quality assessment.

**Findings:** 4,033 citations were identified and 15 were included. No empirical evidence of automated contact tracing’s effectiveness (regarding contacts identified or transmission reduction) was identified. Four of seven included modelling studies suggested that controlling COVID-19 requires high population uptake of automated contact-tracing apps (estimates from 56% to 95%), typically alongside other control measures. Studies of partially-automated contact tracing generally reported more complete contact identification and follow-up, and greater intervention timeliness (0.5-5 hours faster), than previous systems. No meta-analyses were possible.

**Interpretation:** Automated contact tracing has potential to reduce transmission with sufficient population uptake and usage. However, there is an urgent need for well-designed prospective evaluations as no studies provided empirical evidence of its effectiveness.

## Introduction

In response to the rapid global spread of SARS-CoV-2 since December 2019, governments worldwide have applied widespread social distancing measures to attempt to curb transmission^1^. These policies have suppressed case numbers^2,3^ but have substantial economic, social and indirect health consequences,^4^ leading to a growing focus on alternative control strategies.^5^

Contact tracing is a well-established part of infectious disease outbreak management which aims to interrupt chains of infection transmission (for example, through quarantining contacts) and has formed part of many countries’ initial or ongoing response to the COVID-19 pandemic.^6,7^ Traditionally, contact tracing involves a case recalling their recent contacts, who are subsequently contacted and given public health advice to limit onward transmission. A contact tracing system’s ability to reduce disease transmission depends on how rapidly and comprehensively it can identify and (if applicable) quarantine contacts relative to infectious period,^8–10^ and on quarantine adherence.

Typically, contact tracing’s limitations include incomplete recall of contact events by cases, the time taken to notify contacts manually, which can delay quarantine,^11^ and the fact that it is often resource-intensive and time-consuming.^8,9^ Technology could address some of these limitations, including by automating processing of test results or symptom reports^12^ and using smartphones’ capabilities (e.g. Bluetooth) to identify and notify at-risk contacts instantaneously.^11,13^ Automated contact tracing for COVID-19 has been deployed in several countries^14,15^ and is commencing in the UK.^16^ However, the practical, legal and ethical considerations involved are complex^16–18^ and take-up, privacy, security, and testing access have been identified as potential barriers to effectiveness.^17,19^

This rapid systematic review aims to assess the effectiveness of automated and partially-automated contact tracing systems in identifying at-risk contacts and in controlling disease transmission in humans, to inform discussions about the balance between benefits and potential risks of automated contact tracing in controlling COVID-19.

## Methods

This rapid systematic review is registered with PROSPERO (CRD42020179822) and our protocol is available as a pre-print.^20^

### Search strategy and selection criteria

We searched PubMed, EMBASE, and OVID Global Health for articles from any setting published between 1 January 2000 to 14 April 2020. We supplemented this with searches of medRxiv, bioRxiv arXiv, EBSCO Medical COVID Information portal, Cochrane Library and Google Advanced (see Supplementary Information for search terms), and scanned relevant references of included studies. We also included studies identified through professional networks up to 30 April 2020.

Primary outcomes of interest were the number or proportion of contacts identified and the number or proportion of contacts who go on to become cases that are identified (‘contacts’ referring to those considered at-risk due to their exposure to a case). Secondary outcomes included were: impact on either R_0_ or R_e_ (basic or effective reproduction number; the average number of secondary cases infected by one infectious person, in a completely susceptible or real-world population respectively) or other indicators of outbreak control (e.g. completeness or timeliness of contact follow-up or intervention); population uptake (i.e. app uptake or participation); resource requirements (e.g. time, financial resources, testing capacity, training or specific expertise) or cost-effectiveness (e.g. cost per case prevented or per quality-adjusted life year); ethical considerations and lessons learnt from implementation of an automated or semi-automated contact tracing system. Our original protocol included data security, privacy issues and public perception but was modified to exclude these outcomes, partly because they are addressed by the Ada Lovelace Institute report^17^ which was published during our review process, and to focus on evidence of effectiveness from a public health perspective.

We included interventional, observational, modelling and case studies related to automated or partially-automated contact tracing in humans that reported findings regarding at least one outcome of interest. We included studies of COVID-19, SARS, MERS, influenza, or Ebola or, in modelling studies, hypothetical infections spread through respiratory transmission. Studies in which some contact tracing processes were automated (e.g. automated calculation/updating of follow-up periods, contact list generation, alert generation, transmission mapping) but which did not use data from a device as a proxy for contact, or which required users to notify contacts, were considered partially-automated. Purely qualitative study designs were excluded, as were app protocols and studies of monitoring during quarantine. Articles with or without comparators were considered eligible. Both pre-print and peer-reviewed articles were included.

Our search was restricted to full-text manuscripts in English. Non-English language studies flagged for full text review have been collated in supplementary information (table S6). Title and abstract screening was performed by two researchers [IB and TC], with 10% of excluded records dual screened. Full-text screening for eligibility was undertaken by two reviewers [IB and MB/TC]. Discrepancies were resolved by consensus, with an independent view given by a third reviewer [TC or RA]. All exclusion decisions were documented.

### Data analysis

One reviewer [IB] extracted data (see protocol for details)^22^ using a standardised, pilot-tested spreadsheet. Data extraction was reviewed for each study by a second reviewer [MB or TC]. One reviewer quality appraised studies [IB], using the Effective Public Health Practice Project tool for interventional/observational study designs^21^ or using an adapted version of the CHEERS checklist^22^ for modelling studies. In the absence of an appropriate standardised tool for appraisal of descriptive case studies, we documented key factors likely to influence study quality (selection or information bias, confounding, selective reporting, conflicts of interest). We synthesised study findings narratively. We specified in the protocol^20^ that meta-analyses would be considered for ≥three papers investigating a comparable intervention within a similar disease context with a comparable quantitative primary outcome.

### Role of the funding source

There was no specific funding for this project. IB and TC are National Institute for Health Research Academic Clinical Fellows. RWA is supported by a Wellcome Trust Career Development Fellowship [206602].

## Results

We identified 4,033 records from database searches, 398 of which were excluded as duplicates and 110 were reviewed as full text (see PRISMA flowchart in figure 1 for details); two further relevant studies were identified through professional networks and one from reference lists of included studies. 15 records were included and had data extracted. Extracted data are summarised in tables 1-2, which respectively detail key study characteristics including populations, interventions and comparators (table 1) and outcomes and key findings (table 2). Supplementary table S1 details modelling studies’ key assumptions and input parameters and supplementary table S2 details findings and lessons learnt further. We did not undertake any meta-analyses as our pre-specified criteria for this were not met.

**Figure 1:**
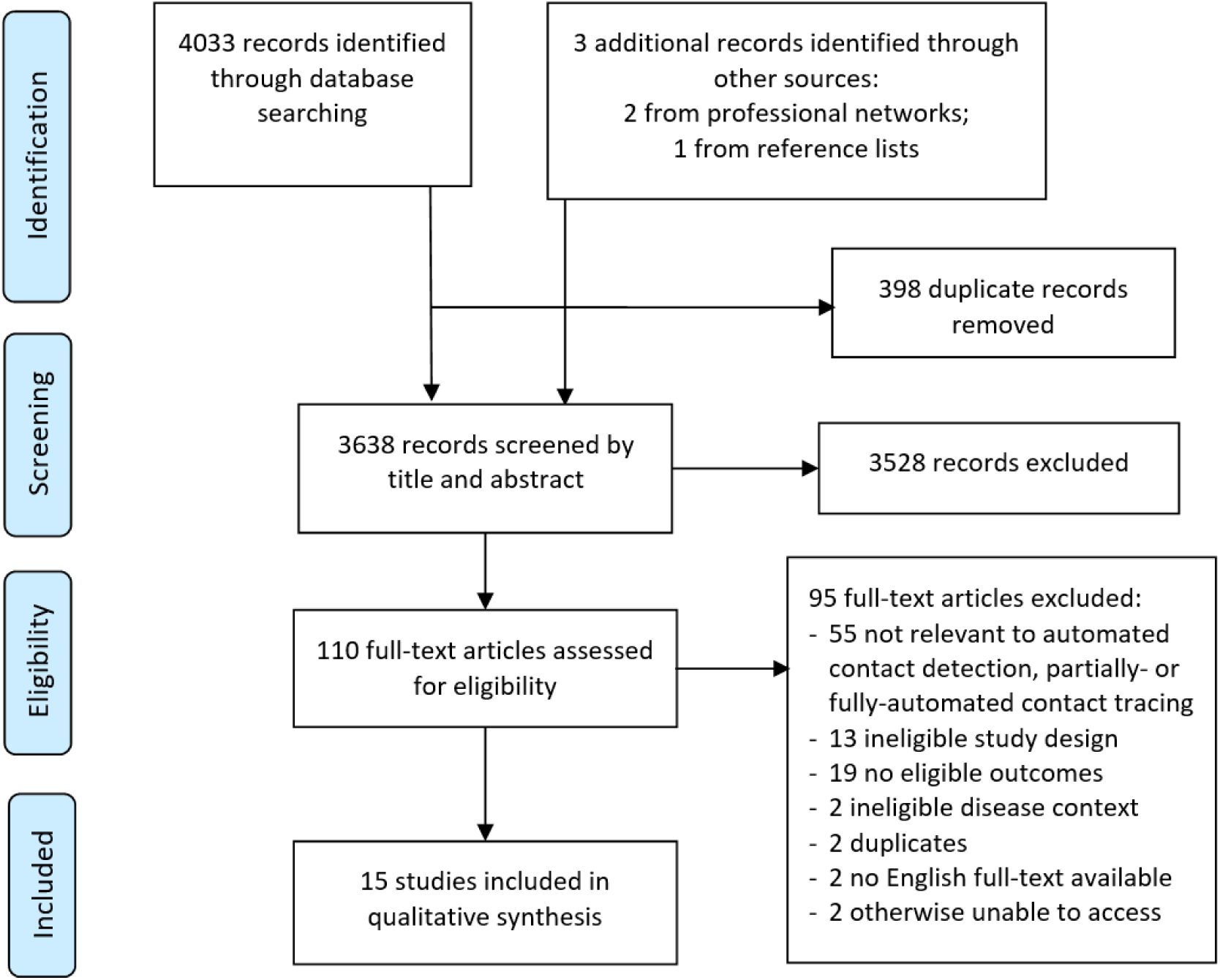
PRISMA Flow diagram (Moher et al. 2009)^48^.

**Table 1:** Summary of study designs, settings, diseases under study and characteristics of populations, interventions and comparators.

| Reference, year | Study design | Summary; Name of app/platform | Study setting - e.g. country, region(s) | Disease(s) under study | Study population details (age range; other identifying characteristics) | Number of participants or app users | Automated (no recall or manual data entry) or semi-automated* | Technologies involved (e.g. GPS, Bluetooth) | Device(s) designed for - e.g. web browser, basic mobile phone, smartphone, multiple | Contact definition used/nature of contacts (e.g. recall-based, proximity-based) | Comparator(s) used |
| --- | --- | --- | --- | --- | --- | --- | --- | --- | --- | --- | --- |
| 1) Studies of fully-automated contact tracing |  |  |  |  |  |  |  |  |  |  |  |
| Bulchanda ni et al. 2020 | Modelling study | Branching-tree mathematical model with derivations of mean-field equations for transitions to 'digital herd immunity' (i.e. $R_0 < 1$ due to automated contact tracing) | N/A | COVID-19 | Hypothetical modelled population | N/A | Automated | Not specified | Not specified | Not specified | None |
| Ferretti et al. 2020 | Modelling study | Modelling study estimating the proportion of transmission from pre-symptomatic individuals (from a series of 40 case pairs), asymptomatic, symptomatic and environmental transmission, and quantifying intervention (case isolation and contact tracing and quarantine) at different delay periods and for different intervention success rates. | N/A | COVID-19 | Hypothetical population network and contact structures, detailed in Fraser et al. (2004)<sup>8</sup> | N/A | Automated | Not specified | Smartphone app (standalone) | Location/proximity-based | Non-automated contact tracing (comparisons based on delay to case isolation and contact quarantine) |
| Hinch et al. 2020 | Modelling study | Modelling study of multiple outcomes under different scenarios involving app-based contact tracing alongside non-targeted NPIs | UK | COVID-19 | Hypothetical population network and contact structures selected to match age-stratified data reported in Mossong et al. (2008).<sup>46</sup> | N/A | Automated | Bluetooth | Smartphone app (standalone) | Proximity-based | Other non-pharmaceutical outbreak control intervention |
| Kim and Paul, 2020 | Modelling study | Mathematical model of automated contact tracing to determine the minimum fraction of the population that needs to participate for it to ensure $R_0 < 1$ . | Not specified | COVID-19 | Hypothetical population | N/A | Automated | Not specified | Not specified | Proximity-based | Not specified |
| Kucharski et al. 2020 | Modelling study | Modelling study of the impact of multiple interventions (including app-based contact tracing, limits on daily contacts and >1 intervention in parallel) on individual-level transmission events | UK | COVID-19 | Hypothetical population with contact structure based on data from the 'BBC Pandemic' study of 40,162 UK participants | N/A | Automated | Not specified | Smartphone app (standalone) | Social contacts (conversational or physical contact), as per the contact definition of the 'BBC Pandemic' study | >1 comparison (including with no contact tracing and manual contact tracing) |
| Xia and Lee, 2020 | Modelling study | Mathematical model of automated proximity-based contact tracing – derivation of formulae to estimate lower and upper bounds on the minimum adoption rate required to achieve $R_0 < 1$ | N/A | COVID-19 | Hypothetical population | N/A | Automated | Proximity-based (further detail not specified); also discusses use of GPS data | Smartphone app or standalone wearable device | Proximity-based | None |
| Yasaka et al. 2020 | Modelling study | Description of TrackCOVID, a decentralised Bluetooth-based contact tracing app including modelling of the population infected at different levels of uptake. | N/A | COVID-19 | Hypothetical population | N/A | Automated | Checkpoints based on QR codes | Smartphone app (standalone) | Face-to-face with manual code scanning | Compared with no contact tracing |
| 2) Studies of partially-automated contact tracing |  |  |  |  |  |  |  |  |  |  |  |
| Danquah et al. 2019 | Proof-of-concept study with phased introduction | Proof-of-concept observational study regarding the introduction of the Ebola Contact Tracing (ECT) app; details number of contacts identified compared with previous system | Sierra Leone | Ebola | Contact tracers and contact tracing coordinators | 86 Contact Tracers, 26 Contact Tracing Coordinators | Partially-automated | Manual data entry | Smartphone app (standalone) | Recall-based | Other (paper-based system) |
| Li et al. 2017 | Case study | Automated identification of contacts within an inpatient setting (lists generated based on user-defined parameters) | Singapore | Multiple including influenza A | Hospital inpatients at Changi General Hospital; system used by IC team | Not specified | Partially-automated | Real-time integration of patient movement and laboratory data | Computer-based infection control management system | Shared room/concurrent contact/duration of contact | Non-automated contact tracing |
| Schafer et al. 2016 | Case study | Case study of app-supported contact tracing using EpiInfo | 7 African countries | Ebola | Used by contact tracers and | No. of contact tracers not | Partially-automated | Manual data entry | Computer-based | Recall-based | Paper-based contact tracing |
|  |  | VHF application; automation of tasks inc. generation of daily follow-up lists; calculation of follow-up window; production of transmission chain diagrams | , 2 US states |  | contact tracing coordinators in 8 countries | specified |  |  |  |  | systems and FIMS (programme developed by WHO) |
| Sacks et al. 2015 | Case study | Case study of the introduction, use and lessons learned from the use of the CommCare contact tracing system in Guinea (c.f. previous paper and Excel-based systems) | Guinea | Ebola | Used by contact tracers | Used by 210 contact tracers (of 366 who were trained) to collectively monitor 9,162 contacts. | Partially-automated | Manual data entry | Smartphone app (standalone) | Recall-based | Paper-based contact tracing system (used in parallel within Guinea by other contact tracers) |
| Tom-Aba et al. 2015 | Case study | Case study of the use of an app (ODK Collect app) developed using OpenDataKit and FormHub to support contact tracing in Nigeria. Automated alerting and SMS if any contact met the probable case definition. Also refers to Ebola Sense app (supports follow-up of identified contacts, includes automated search functionality to assign contact tracers). | Nigeria | Ebola | Used by contact tracers | Not specified; all public health personnel carrying out contact-tracing for Ebola cases | Partially-automated | GPS (for accountability of contact tracers rather than directly within tracing efforts) | Smartphone (or tablet) app (standalone) | Recall-based | Paper-based contact tracing systems (data manually entered onto a single computer) |
| 3) Studies of automated contact detection in a relevant disease context (without subsequent contact tracing or contact notification) |  |  |  |  |  |  |  |  |  |  |  |
| Aiello et al. 2016 (iEpi sub-study) | Sub-study (descriptive observational study) within a cluster-RCT | 'iEpi' sub-study within a university-based trial; participants given smartphones which were used to detect other study devices as well as nearby Bluetooth-enables devices to map proximity contacts | US | Influenza | Students aged ≥18 | 103 (iEpi sub-study) | Automated contact detection only | Bluetooth | Smartphone app (standalone) | Proximity-based | None |
| Al Qathrady et al. 2016 | Observational and modelling (simulation) study | Simulation of disease spread and contact tracing using a contact network recreated from WiFi traces; explores different approaches to prioritising investigation/follow-up of at-risk contacts | Not specified | Hospital-based outbreaks | University faculty and students (contact data based on Wifi use on a university campus across 6 buildings) | Wifi trace data from 34,225 users of 6 University buildings | Automated contact detection only | WiFi network traces | Mobile devices (further detail not specified) | Proximity-based | None |
| Voirin et al. 2015 | Proof-of-concept observational study | Observational pilot study combining micro-contact data from wearable proximity sensors and virological data to investigate influenza transmission within an elderly care unit of a hospital | France | Influenza A and B | Median age (y): 24 - doctors; 30 - nurses; 89 - patients. % female: Doctors - 67%, Nurses - 78%, Patients - 73% | Total 84 (32 nurses, 15 doctors, 37 patients), of 48 healthcare workers and 44 total patients (86% participation rate) | Automated contact detection only | Radio-frequency identification (RFID) | RFID proximity sensors | Proximity-based | None |

\*Semi-automated: requires some manual recall by cases of their contacts, manual data-entry into an electronic system with some automated features and/or involvement in informing contacts
**Abbreviations and acronyms:** FIMS: Field Information Management System; QR code: Quick Response code; RCT: Randomised Controlled Trial; RFID (Radio-frequency identification); SORMAS: Surveillance and Outbreak Response Management and Analysis System; US: United States; UK: United Kingdom; WHO: World Health Organization.

**Table 2:**
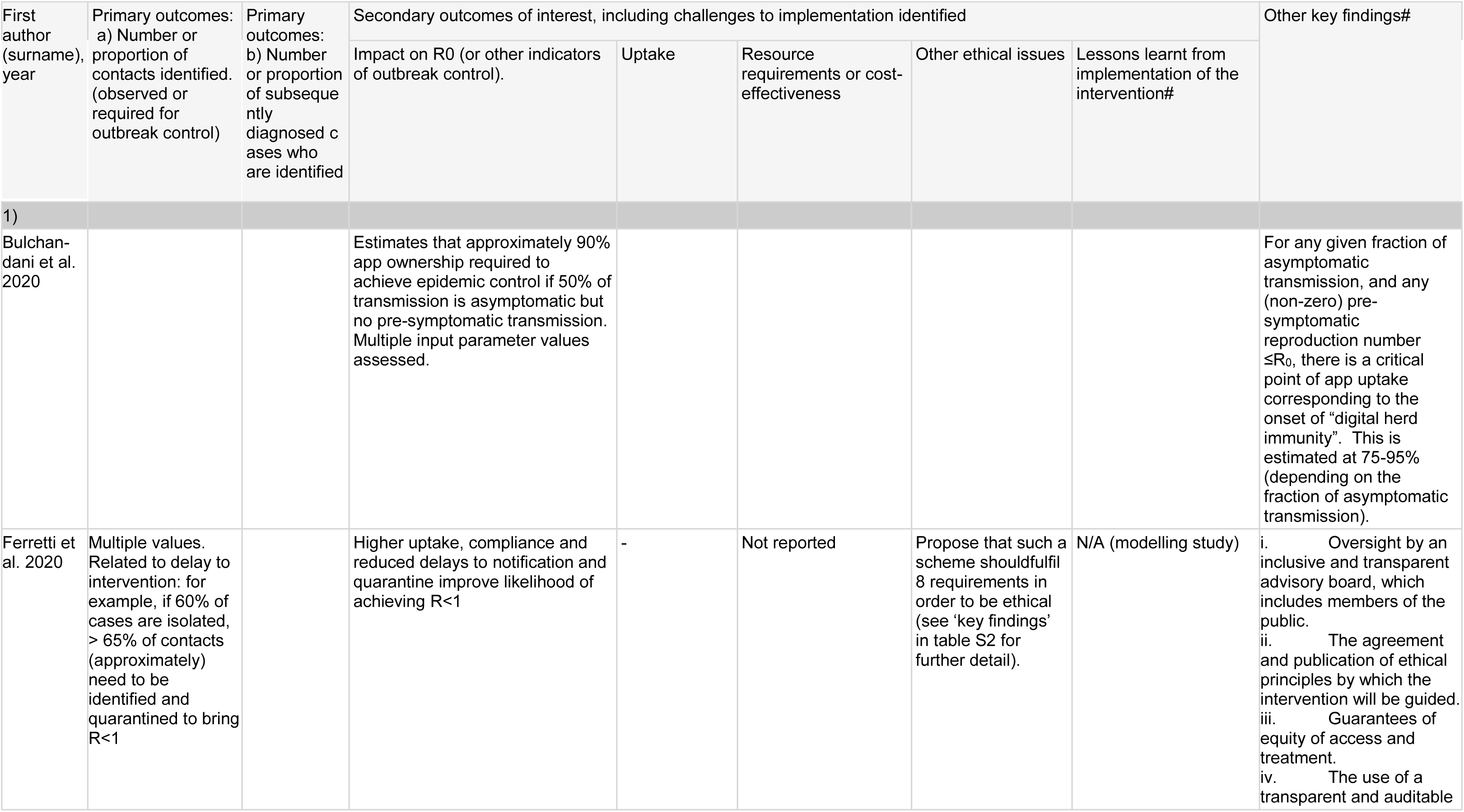
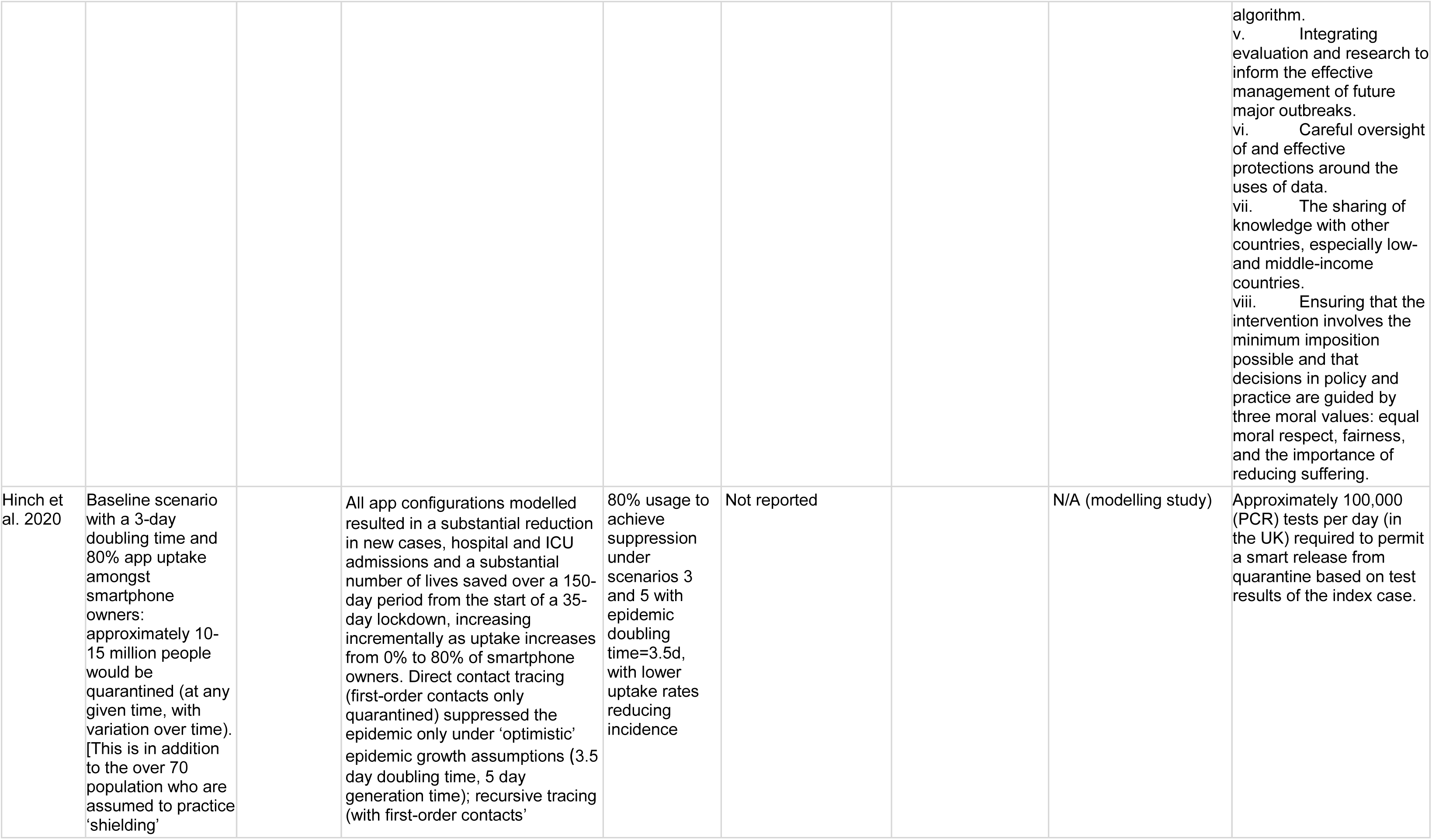
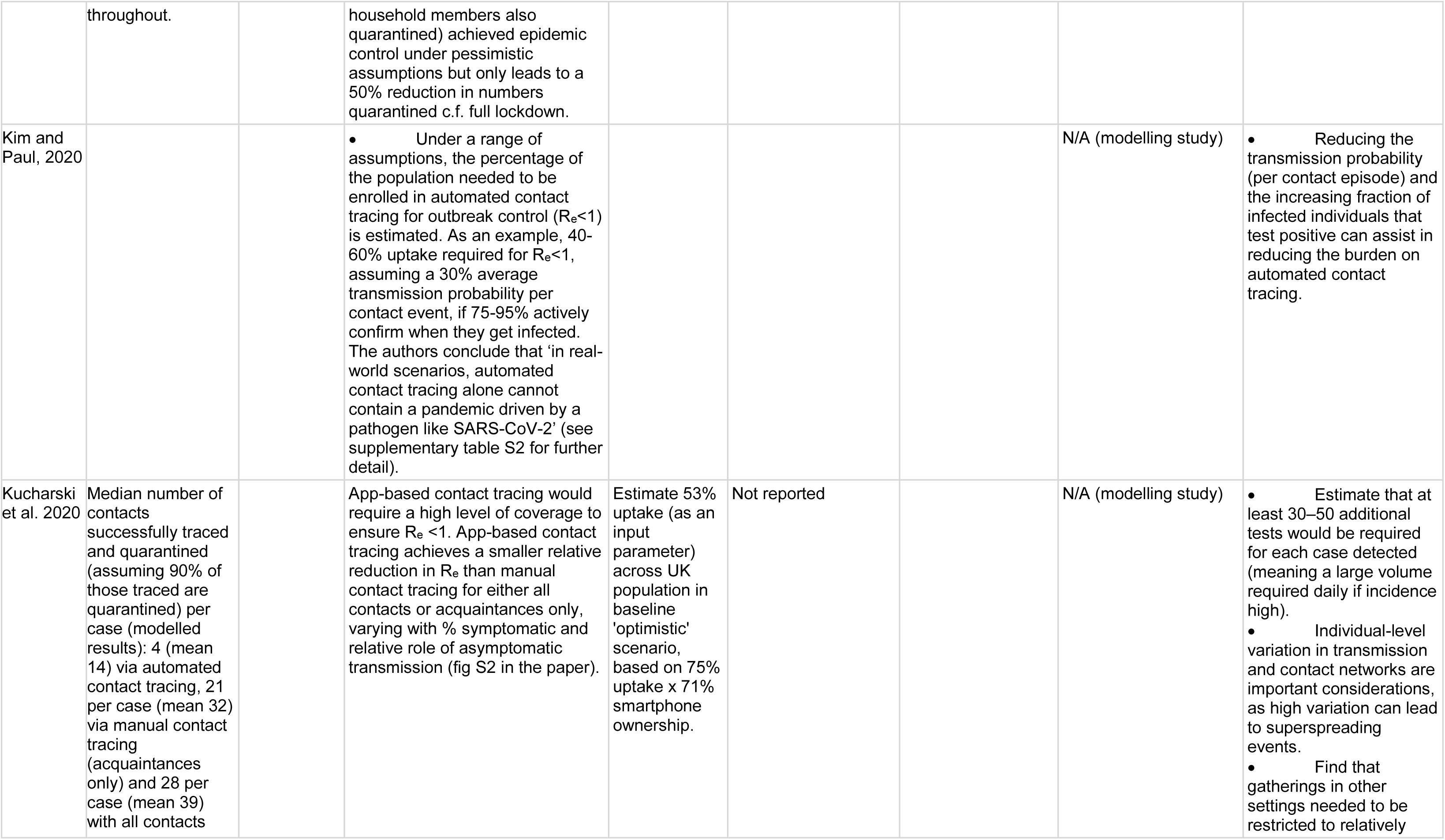
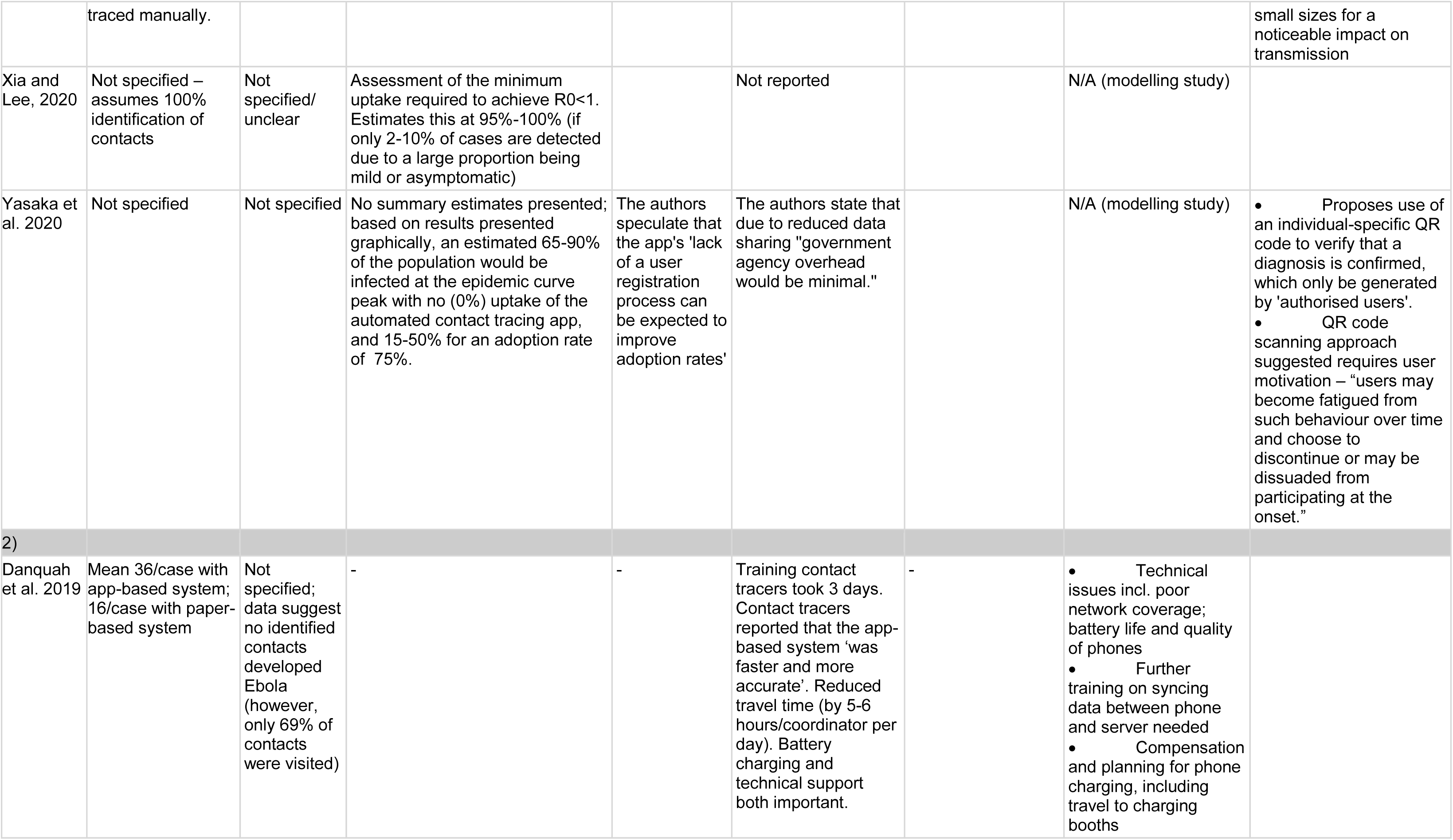
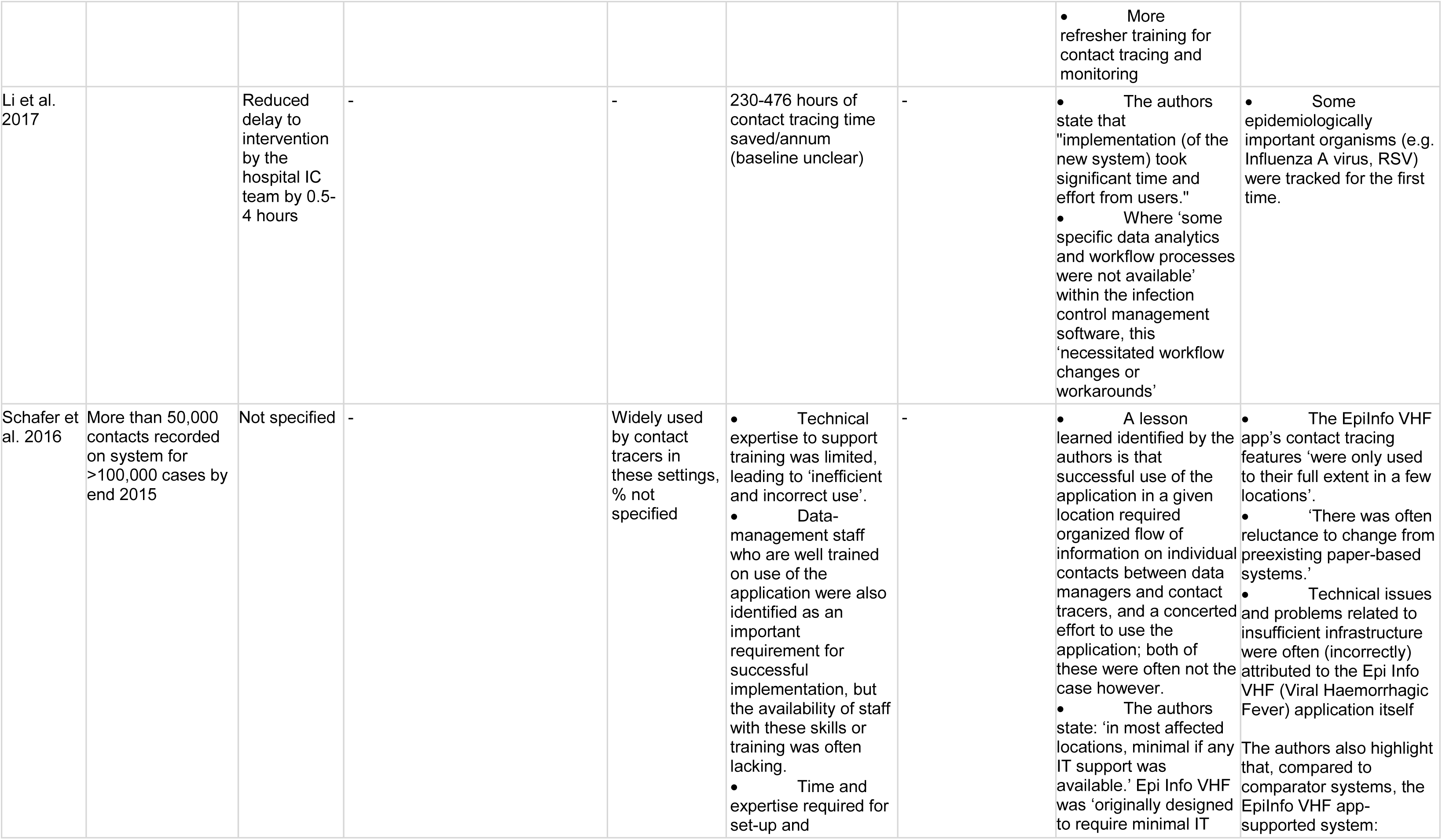
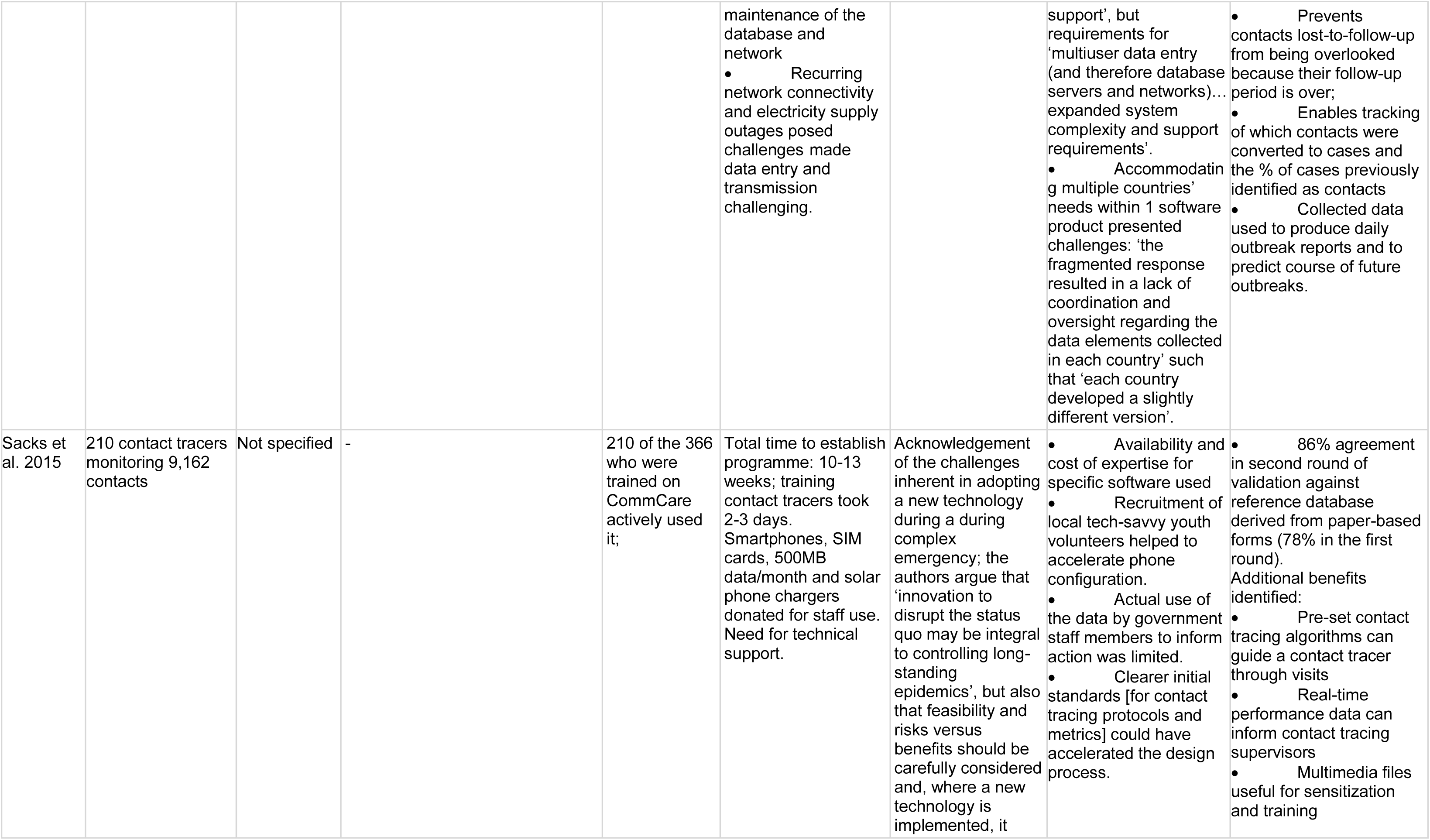
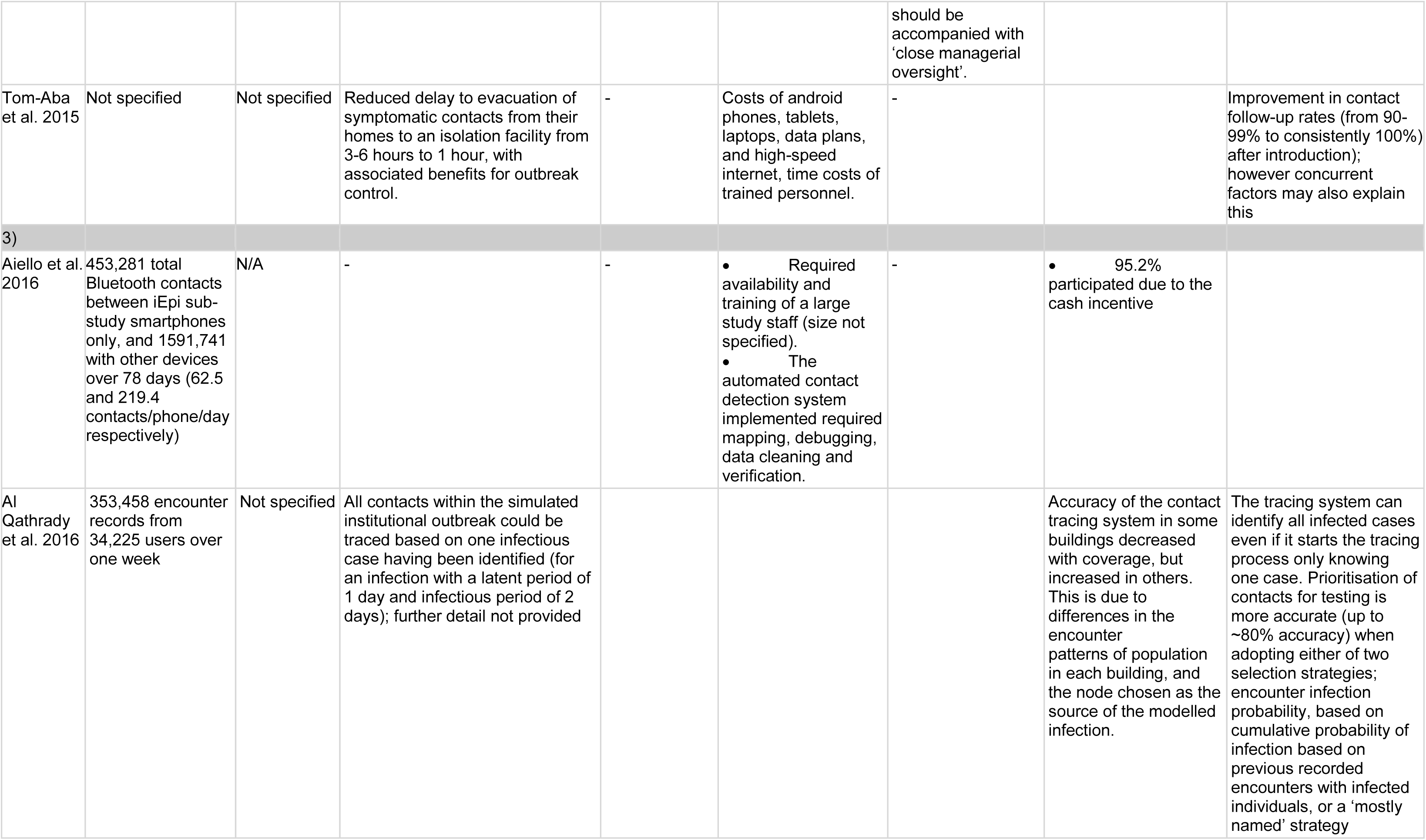
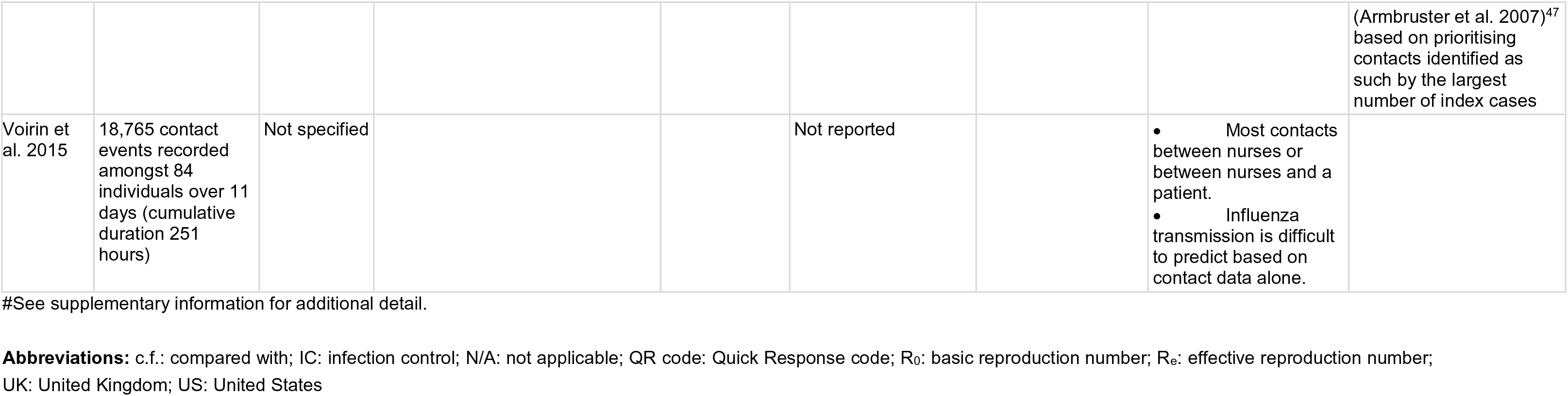
Summary of outcomes and other key findings.

Included studies’ findings are detailed below in three categories, the third of which was defined post-hoc: (1) seven studies that addressed automated contact tracing directly (all modelling studies focused on COVID-19)^11,23–28^; (2) five descriptive observational or case studies of partially-automated contact tracing (four studies related to Ebola^29–32^ and one hospital infection control system)^33^, and (3) three studies of automated contact detection within a relevant disease context but without subsequent tracing or contact notification.^34–36^ No study assessed ethical considerations relevant to decision-making were discussed by two studies (see table 2 for details).^11,29^

### 1. Studies of automated contact tracing

We found seven studies of automated contact tracing; all were mathematical modelling studies, with varied assumptions (supplementary table S1). Five of these addressed smartphone apps^11,23–26^ specifically, alongside other wearable devices in one study.^25^ Two studies related to an unspecified type of device carried by users.^29,30^ No studies contained data on our primary outcomes using the same definition used in our protocol (number or proportion of contacts (and of contacts that go on to become cases) identified); however, two of the seven modelling studies provided data of a comparable and relevant nature in the form of estimated numbers of contacts quarantined.

In a modelling study of control measures for COVID-19 in the UK, Kucharski and colleagues^23^ estimate that a median of four contacts per case (mean 14) would be quarantined under automated contact-tracing, compared to 28 (mean 39) with all contacts traced manually, assuming 90% adherence to quarantine. Also modelling COVID-19 in the UK, Hinch and colleagues^24^ assumed 100% initial adherence to quarantine and 80% uptake amongst smartphone owners, and estimated that approximately 10-15 million people would be quarantined (cumulatively, at any given time, alongside the over 70 ʻshielding’ population), but did not present numbers of contacts identified per case. Three studies described an approximately quadratic relationship between population uptake of an automated contact tracing tool/app and associated reductions in transmission,^27,28,23^ such that 80% uptake might enable notification of approximately 64% of the contacts who would be notified in an optimal contact tracing system; with 50% uptake the corresponding figure is 25%.^28^

Only Kucharski and colleagues^23^ directly compared automated and manual contact tracing’s modelled impacts on R_0_ or R_e_. Under ʻoptimistic’ assumptions including 75% uptake amongst smartphone owners (see supplementary table S1), and assuming equal maximum delays to quarantine of contacts under automated and manual scenarios, they estimated that automated tracing alone reduced R_e_ by 44% whereas manual tracing of all contacts reduced it by 61%. Hinch and colleagues^24^ did not compare automated and manual contact tracing or report impacts on R_e_. Both studies^23,24^ found that suppressing the COVID-19 outbreak required concurrent measures (e.g. shielding vulnerable groups,^24^ remote working and limits on numbers of contacts per day^23^) alongside automated contact tracing. Most scenarios modelled by Hinch and colleagues^24^ did not achieve containment (equivalent to R_e_<1), except when quarantining all household members of contacts who had direct contact with a case.^24^

Two other modelling studies of automated contact tracing for COVID-19 reported similar findings: one study^27^ estimated 75%-95%, and another 90-95%,^25^ population-wide uptake to be required to bring R_e_ below 1. Several studies found that, even below this threshold, increasing uptake was associated with reduced COVID-19 incidence.^27,24,26^

Regarding resource requirements, one study^26^ estimated that approximately 200,000 tests/day would be required for test-based quarantine release in the UK; another^23^ estimated 30-50 tests required per case detected. No other secondary outcome data were reported for this section.

### 2. Studies of partially-automated contact tracing

We found a total of five studies of partially-automated contact tracing, which all automated some processes within systems involving human contact tracers or infection control staff. Li and colleagues^33^ profiled a hospital-based system which automatically alerts staff to new infections by target organisms and generates contact lists using user-defined parameters (e.g. having shared a room, concurrent contact, duration of contact). Four studies^29–32^ focused on software applications used to manage Ebola outbreaks.

Three of these studies reported data relevant to our primary outcomes; in one^32^ a mean of 36 contacts per Ebola case were recorded for cases where contact tracers used an app (‘Ebola Contact Tracing application’), compared with 16 per case under the pre-existing paper- and Excel-based system. In a second^30^ study >100,000 investigated cases and >50,000 contacts were recorded in the Epi Info Viral Hemorrhagic Fever (VHF) application by contact tracers across 7 African countries and 2 US states by end 2015; the reason for this apparent low ratio of only approximately 0·5 contact per case recorded was unclear. A third^29^ study of the CommCare app, a partially-automated application with algorithm-based decision-support features (e.g. prompting referral for testing on entry of data indicating that a contact developed symptoms) and which updated a data visualisation dashboard automatically every hour, reported 9,162 contacts but the number of cases of these contacts was unspecified. No other primary outcomes were reported in these studies.

Contact follow-up rates were increased compared with previous paper-based systems in two studies in this section.^31,32^ Two studies reported improved intervention timeliness (e.g. quarantine or isolation) compared with previous, non-automated systems; by 2-5 hours in one study^31^ and by 0·5-4 hours in another.^33^

Three studies^31,33,34^ detailed the hardware, software and supporting infrastructure requirements of partially-automated contact tracing systems; these included smartphones, tablets, laptops, SIM cards, data plans, high-speed internet and phone battery charging. No study in this section provided cost information and only one^29^ detailed implementation duration (10-13 weeks). Li and colleagues^33^ reported approximately 230-476 hours/year of contact tracing work was saved by a partially-automated infection control management system in one hospital. In another^32^ contact tracers reported that the app-based system ʻwas faster and more accurate’ and eliminated substantial travel time (5-6 hours per coordinator daily). Technical support needs, including for training, were a recurrent theme. For example, one study^30^ stated that training ʻwas often provided by staff who had received only minimal training themselves’ leading to ʻinefficient and incorrect use’; technical expertise was highlighted as an important but limited resource in two others.^31,32^ One study^29^ reported that training contact tracers took 2-3 days and another^32^ 3 days.

Lessons learnt included the importance of reliable internet and electricity infrastructure,^30,32^ and the value of customising systems based on local priorities.^30^

### 3. Other studies relevant to automated contact tracing

We found three studies of contact detection in a relevant disease context but without subsequent tracing or contact notification: one studied students’ smartphone contact patterns^34^; another integrated radio-frequency contact and virological data^35^ and another used wifi traces to model a hypothetical epidemic.^36^

None detailed a primary outcome precisely as specified; however, participants in one study^34^ averaged 219 contacts/day with devices of any kind, whilst another^35^ observed 18,765 contact events amongst 84 participants over 11 days (but only 4 influenza transmission events). One study^34^ referred to the need for availability and training of a large study staff. Lessons learnt are detailed in table 2, with further detail in supplementary table S2.^36,37^ No other secondary outcomes were detailed.

#### Quality assessment

Study quality was variable and quality assessments are detailed in supplementary tables S3-5. The quality of studies in categories 2 and 3 was generally limited by their observational and often descriptive nature, without pre-specified protocols (except in one,^32^ where this was modified during the study). Many were subject to possible confounding, selection bias and selective reporting. Amongst the modelling studies, some (e.g. Ferretti et al.,^11^ Hinch et al.^24^ and Kucharski et al.^23^) included detailed methods, conducted a range of sensitivity analyses (except one)^11^ and provided their model code. Others provided limited justification of the model structures or assumptions used and did not account for uncertainties or conducted only limited sensitivity analyses.

## Discussion

We found no epidemiological studies comparing automated to manual contact tracing systems and their effectiveness in identifying contacts. The modelling studies we identified found that automated contact tracing’s effectiveness depends on both population uptake (e.g. of contact tracing apps) and timeliness of intervention (e.g. quarantining contacts).^11^ Uptake is particularly important since both infectious cases and their contacts need to have and be using a system for it to have any effect, leading to a quadratic relationship such that effectiveness drops off steeply as participation falls. Even under optimistic assumptions (e.g. 75-80% app uptake amongst smartphone owners and 90-100% adherence to quarantine), automated contact tracing appears unlikely to achieve control of COVID-19 without concurrent measures;^23–25^ this is even more the case in settings with low smartphone ownership.^27^

Strengths of this review include the comprehensive search strategy and pre-specified eligibility criteria and screening process. With its focus on outbreak control, it also addresses timely, policy-relevant questions. We quality-assessed all studies, but were unable to undertake meta-analysis and formal assessment of publication bias. Other limitations include the lack of eligible empirical studies of fully-automated contact tracing and a paucity of evidence related to ethical concerns or cost-effectiveness. The modelling studies reflect substantial uncertainty; for example, if environmental transmission (e.g. via droplet contamination of surfaces) of COVID-19 occurs more often than typically assumed by these studies, this would undermine their validity. Given these uncertainties, which relate both to SARS-CoV-2’s transmission and epidemiology and to human behaviour under new, untested scenarios, it is difficult to objectively appraise how realistic the modelling studies’ assumptions (and therefore results) are. Additionally, our review was limited to English-language studies due to short timescales.

Our primary outcomes, regarding numbers and proportions of contacts (including of those who become cases) identified, are a key gap in current evidence, and important metrics for evaluation. The integration and relative impacts of manual and automated systems run in parallel are also unexamined. Pre-symptomatic transmission may be substantial in COVID-19,^37,38^ making timeliness of quarantine critical.^8,11^ However, the relative timeliness of automated versus manual contact tracing systems is unknown, though partially-automated systems appeared to reduce delays to quarantine.^31,33^ Additionally, whether quarantine adherence differs between automated and manual systems is unknown. Automated notification might be psychologically different from receiving a phone call from a human contact tracer who can give detailed information about what action to take and why, check understanding and address questions or concerns. A previous review^39^ found adherence to be extremely variable and influenced by multiple factors, including risk perception and social and financial protections.

Academics have recently warned of automated contact tracing’s risks including ʻmission creep’ towards unprecedented surveillance, and eroded public trust should data be misused or hacked.^40^ These are clearly important considerations,^19,41,42^ although beyond our review’s scope. Trade-offs between privacy and utility are discussed elsewhere^13,19^ and may vary between system architectures,^43^ particularly ʻcentralised’ systems, which involve data being uploaded to a central server, and ʻdecentralised’ systems, which are more strongly privacy-preserving, keeping co-location data on users’ phones. Decentralised systems also benefit from Apple and Google’s support.^16^ However, Fraser and colleagues^43^ find that centralised systems assess transmission risk more accurately (reducing numbers quarantined), enable better optimisation, are less susceptible to false reports, and more readily evaluated.

Optimising risk thresholds in order to minimise transmission risk and numbers quarantined simultaneously is a key challenge for any contact tracing system,^10^ particularly in view of quarantine’s adverse psychological impacts^44^ and wider harms^4^. However, this relies on gathering and analysing high-quality data. Where automated contact tracing systems are deployed, they should be evaluated rigorously,^11,43^ including through prospective cohort studies and qualitative studies.

Wider concerns around digital exclusion and broader ethical concerns have been highlighted elsewhere^18,45^ including in the Ada Lovelace Institute review^17^. Some particularly vulnerable populations (e.g. older and homeless people) are also less likely to own a smartphone, potentially amplifying their risks.^18,13^ Such challenges are more acute still in low-income countries.^27^

Given substantial remaining uncertainties about automated contact tracing systems’ effectiveness, large-scale manual contact tracing is likely to be required to control COVID-19, alongside automated approaches and other measures such as remote working by a proportion of the population and limiting daily social contacts. Moderate uptake of automated systems could contribute to reducing transmission and could offset some of the work of manual contact tracing. However, such benefits must be weighed against implementation costs and broader risks. Decision-makers should use all available evidence to ensure that contact tracing systems are as effective, equitable and acceptable as possible, and that they are deployed within integrated outbreak responses.

## Data Availability

The data supporting this systematic review are from previously reported studies, which have been cited. The processed data supporting the findings of this study are available within the article and/or its supplementary material.

## Contributor statement

IB and RWA developed the concept of the review. All authors contributed to the development of the study protocol. Title and abstract screening was performed by two researchers [IB and TC], with 10% of excluded records checked in duplicate. Full-text screening for eligibility was undertaken by two reviewers [IB and MB or TC]. RWA gave an independent view in case of any discrepancies. IB extracted data; this was verified by TC and MB. IB quality appraised studies and produced tables and figure 1. IB wrote the first draft of the manuscript with input from TC, MB and RWA; all authors contributed to drafting and editing the manuscript.

## Competing interests

We declare no conflicts of interest.

## Funding

There is no specific funding for this project. IB and TC are NIHR (National Institute for Health Research) Academic Clinical Fellows. RWA is supported by a Wellcome Trust Career Development Fellowship [206602].

## Notes

### Competing Interest Statement

The authors have declared no competing interest.

### Clinical Protocols

https://www.medrxiv.org/content/10.1101/2020.04.14.20063636v1

### Author Declarations

Ethical approval and patient consent are not required as this is a systematic review using already published (peer-reviewed and pre-print) articles; we did not access or use any individual patient data for this study.

